# The incidence and prevalence of Dupuytren’s disease in primary care: Results from a text mining approach on registration data

**DOI:** 10.1101/2024.07.10.24310207

**Authors:** Roel J. M. van Straalen, Michiel R. de Boer, Francine Vos, Paul M. N. Werker, Dieuwke C. Broekstra

## Abstract

**Background:** The focus of research and management of Dupuytren’s disease (DD) is shifting from relieving symptoms in the later stages of disease towards the prevention of contractures. Treatment services might likewise shift towards primary care. Studying characteristics of DD patients who seek medical care for the first time, may identify a symptomatic target group for early DD treatments. We present the first study that estimates the incidence and prevalence of DD in primary care by applying a text-mining algorithm to registration data.

**Methods:** This is a population-based cohort study using electronic health records from Dutch general practices involved in a regional research network. Descriptive statistics were used to describe sex, age, comorbidities and lifestyle factors, the latter two were identified via International Classification of Primary Care (ICPC) codes. Incidence rate was calculated as number of patients with a first contact for DD/1000 person years for the years 2017 to 2021, point prevalence as the percentage of patients with a contact for DD in 2021. DD contacts were identified using a text-mining algorithm.

**Results:** The incidence ranged between 1.41 to 1.72/1000 person years and the overall prevalence was 1.99%. Incidence and prevalence are higher among males and increase with age, peaking between 61 to 80 years.

**Conclusions:** Our results of prevalence and incidence of DD in primary care give an insight into the relevant population of patients with symptomatic DD that might be the future target group for potential disease controlling treatments.

## Introduction

Dupuytren’s disease (DD) is a fibrotic disorder of the hand and is characterized in the early-stages of the disease by formation of nodules or pits in the palm of the hand, which eventually may develop into cords that can bend the fingers. These contractures often occur in a later stage of disease and may lead to functional limitations that can interfere with daily activities.(1,2) The course of disease varies. A retrospective study showed that 50 percent of patients developed progressive flexion deformities during a ten year follow up.(3) Other prospective studies reported progression in 21–50 percent of patients within seven to eighteen years.(4,5)The main treatment in patients having pronounced impairment because of flexion deformities is surgical division or excision of the cords. Surgical treatment is symptomatic and unfortunately does not cure the disease and recurrences may develop, necessitating revision treatment. (6)

The focus of DD research and management is shifting from relieving symptoms in the later stages of disease towards the prevention of contractures. Interventions such as corticosteroid or adalumimab injections, or radiotherapy, which aim to slow down or stop progression of early-stage DD, have been studied.(7) A recent randomized controlled trial reported that repeated intranodular adalimumab injections resulted in softening and size reduction of early-stage DD nodules, which continued to decrease further after the final injection.(8) The ideal target group for this disease controlling treatment consists of patients who present with early DD symptoms, but are prone to progress. Therefore it is important to map patients who seek medical care for DD for the first time. Usually, these patients first present in primary care. In many European countries and part of the health plans in the United States, a general practitioner (GP) or family doctor acts as a gatekeeper and determines whether or not patients require secondary care.(9)

Since primary care is less expensive and more accessible than secondary, specialized health care, the decision for referral is a vital component of demand management and health care costs.(10,11) Therefore, if possible, treatment services should shift towards primary care in the future.

There have been many studies on the prevalence of DD in the general population.(12–17) However, the majority of people with DD in the general population have asymptomatic nodules without limitations (12) and are probably less likely to seek medical attention.

Studying the frequency of symptomatic DD patients who seek medical care will identify the number of patients in the target population that could be eligible for early DD treatment. Patients registries provide unique opportunities to study the incidence and prevalence of conditions. So far, one study reported the prevalence and incidence of DD patients presenting in primary care using registration data, based on codes registered by GPs.(17) However, this may lead to misclassification or underestimation, because of coding errors.(18)

This study therefore aims to assess the incidence and prevalence of DD in Dutch general practices by analyzing registration based data using codes and an additional text-mining algorithm.

## Methods

### Study design

This is a registration-based study in a dynamic population of patients from general practices in the Northern part of the Netherlands. In the Netherlands, GP’s act as gatekeepers in the health care system, and thus present the point of entry of all DD patients into the health care system.

### Study population and setting

We used primary care data from the general practitioner-registration database (“Academisch Huisarts Ontwikkel Netwerk”, AHON) of the Northern region of the Netherlands. It comprises care data of 58 participating general practices including data from around 500.000 patient records. The registry currently accounts for 10.5% of the population in the North.(19) General practices provide patient data from 5 years prior to the start of the registration at AHON. Diagnoses are recorded according to the International Classification of Primary Care (ICPC); symptoms, treatment policies and referrals are recorded in free text. (20,21)

The data is stored and a trusted third party ensures pseudonymisation of patient-identifying data. Patients have the possibility to object to having their data included in the database. For the current study, data up to and including the 31^st^ of December 2021 was extracted. A patient’s registration period started from registration, and ended either by the patient’s death, registration termination, the end of data collection from their practice or the end of the study period (31-12-2021).

Studies using this database do not fall within the scope of the Medical Research Involving Human Subjects Act (WMO) according to the medical ethical committee of the UMCG (research register number 202100077), and therefore require no further ethical approval. We did obtain approval of the scientific committee of the AHON (ID number 74).

### Participants

Participants over 18 years and up to 105 years were selected. Participants with missing registration dates, registration dates prior to date of birth or deregistration dates past 2022 were seen as coding errors and were therefore excluded. The patient-identifying data for pseudonymisation included date of birth, gender and postal codes. If participants moved, or moved from one practice to another during the study period, a duplicate participant could occur (i.e. different ID numbers belonging to the same participant). Duplicate participants were identified via two participant identification pseudonyms and listed registration fees. Subsequently part of the data belonging to the same patient was either deleted or data was merged to avoid duplicates (*Supplementary data: appendix 1)*.

**Figure 1.**
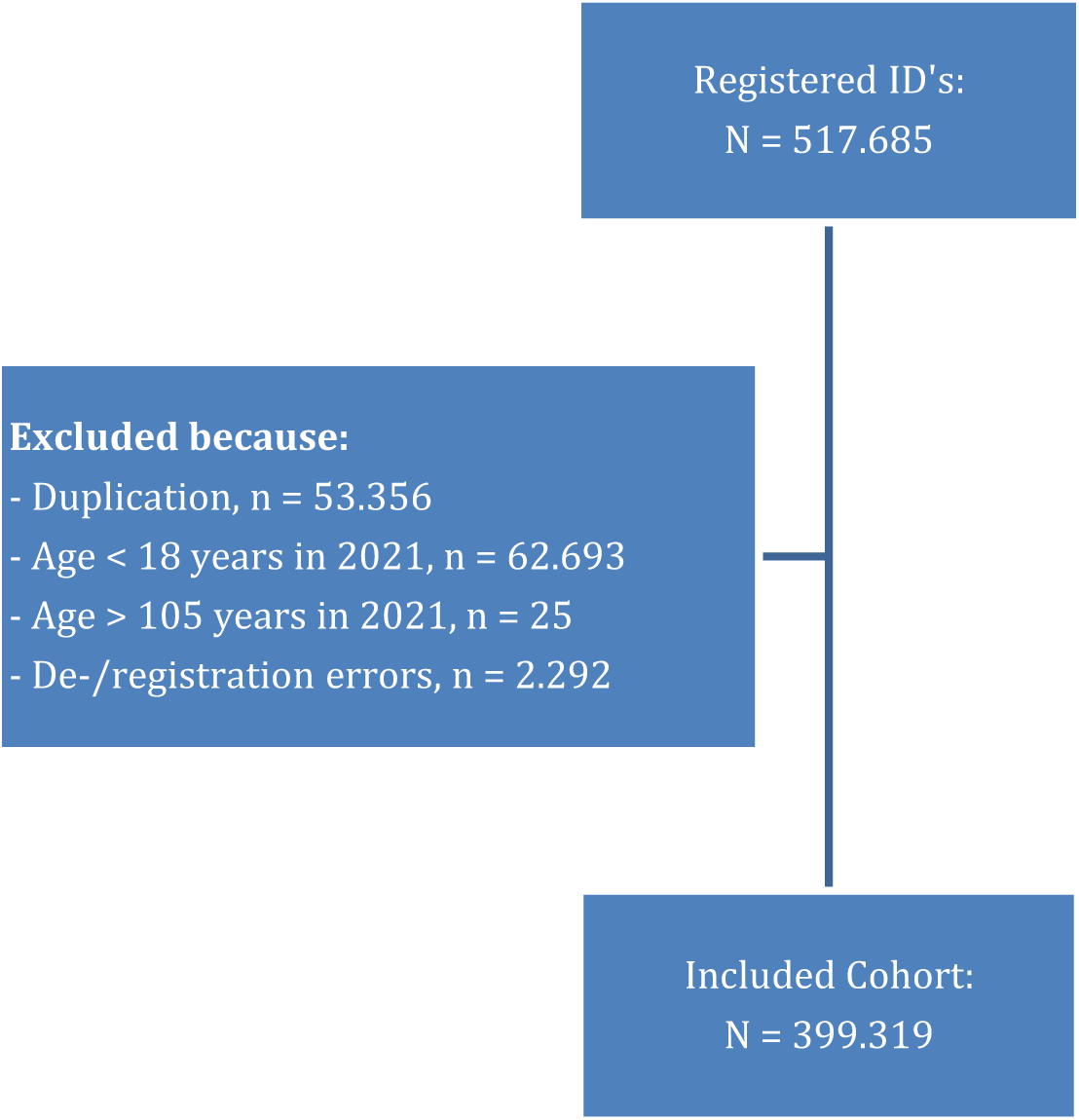
Number of registered person IDs and final participants from the AHON registry up to and including 31st of December 2021.

### Data extraction

Firstly, we extracted all contacts with ICPC code L99.03 (“Dupuytren’s contracture”) or with the letters “Dup” in the free text. Secondly, we extracted three variables based on these contacts: DD diagnosis, the date of first contact for DD and the number of contacts for DD per patient.

DD diagnosis was defined as a contact with ICPC code L99.03 or as a contact with DD mentioned by the GP in free text of the contact. Due to the large number of contacts, we created a text-mining algorithm to determine the diagnosis DD from the free text fields (*Supplementary data: appendix 2)*. The validation of this algorithm was done in three steps:

1. Authors RvS and DB manually evaluated a random sample of 5% of all extracted contacts independent from each other and decided on the DD diagnosis based on the free text. Their scores were compared and an agreement of 0.95 (Cohen’s Kappa) was found.
2. The text-mining algorithm was run on the same sample and adjusted until the agreement between the algorithm scores and the scores from the joined judgment from both authors was 0.95 for DD diagnosis.
3. Finally, the text-mining algorithm was run on a new random sample of 2.5% of the total cohort for validation. The scores were compared to the scores of the first author and had an agreement of 0.93 for DD diagnosis.

Newly diagnosed patients were identified as those having their first contact for DD, without prior DD consultations. The date of first DD contact was established as the earliest contact where DD was diagnosed.

We extracted the sex and age of all participants registered. The following Dupuytren correlates were also extracted based on their ICPC codes: diabetes mellitus, excessive smoking, alcohol abuse, hypercholesterolemia, obesity and epilepsy.

### Statistical analysis

All statistical analyses were executed using R (version 4.0.5). Descriptive statistics of the total cohort and of patients with DD were presented by numbers, percentages, and median and interquartile range (IQR).

We determined the incidence of DD for the years 2017 to 2021, stratified by gender and age categories, by creating sub-cohorts for each year in which participants who were not at risk were excluded. Participants were not at risk when they 1) entered the AHON database later then the year of analysis, 2) left the AHON database prior to the year of analysis, and 3) had a DD diagnosis prior to the year of analysis.

We calculated the incidence for each created subcohort by dividing the number of newly diagnosed DD patients that year by the person-years at risk.

We calculated the 2021 mid-year point prevalence of DD stratified by sex and age categories (age <40, 40-50, 50-60, 60-70, 80-90 and 90+) of participants registered in 2021. The number of DD patients in 2021 was divided by the number of participants registered at the 1st of July 2021 (mid-year population). Confidence intervals were calculated using the package *tidyverse* and function ‘binom.exact’.(22) We also calculated an overall age standardized prevalence using the method of direct standardization with function ‘ageadjust.direct’ in package *epitools*.(23,24).

## Results

### Baseline characteristics

Within our cohort of 399,319 participants we identified 3,361 patients with DD of which 60.7% was male. The median age at diagnosis was 64.5 years (IQR 56.9 – 72.1). As shown in table 1, the median age was higher among DD patients compared to the overall population, DD patients were more often of male sex and there was a higher percentage of relevant comorbidities among DD patients.

**Table 1.** Mid-year characteristics of the population-based cohort from the AHON registry in 2021.

|  | **Overall population** | **DD patients** |
| --- | --- | --- |
|  | **(n = 226,503, %)** | **(n = 2,826, %)** |
| Age (mediaan, IQR) | 52.4 (35.3 – 67.1) | 68.8 (60.8 – 75.9) |
| Males (n, %) | 112,142 (49.5) | 1,700 (60.2) |
| Diabetes Mellitus, yes (n, %) | 17,746 (7.8) | 550 (19.5) |
| Excessive smoking, yes (n, %) | 17,693 (7.8) | 288 (10.2) |
| Alcohol abuse, yes (n, %) | 2,583 (1.1) | 76 (2.7) |
| Hypercholesterolemia, yes (n, %) | 21,712 (9.6) | 533 (18.9) |
| Overweight/Obesity, yes (n, %) | 15,041 (6.6) | 212 (7.5) |
| Epilepsy, yes (n, %) | 2,899 (1.3) | 55 (2) |

### Incidence

Figure 2 shows that the incidence rates were fairly stable over time ranging from 1.65 (95%CI 1.41 – 1.91) to 2.08 (95%CI 1.82 – 2.39)/1000 person years for males and from 1.12 (95%CI 0.93 -1.33) to 1.36 (95%CI 1.15 – 1.60)/1000 person years for females. The overall incidence rates ranged from 1.41 (95%CI 1.26 – 1.58)/1000 person years in 2020 to 1.72 (95%CI 1.55 – 1.91)/1000 person years in 2019.

**Figure 2.**
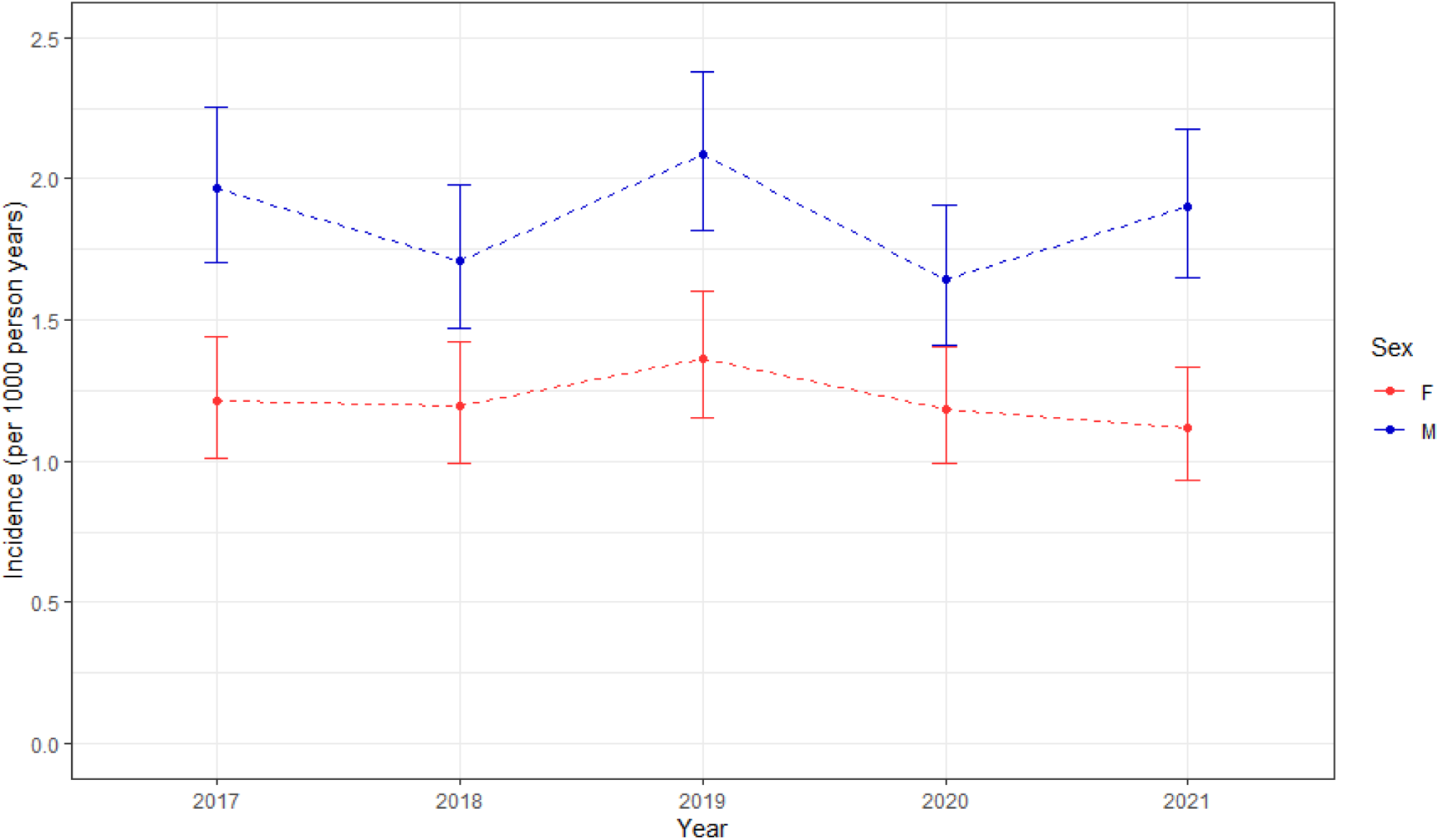
The incidence of Dupuytren’s disease in the AHON registry specified for sex and year of observation.

Overall, the incidence was higher for males than for females and the highest incidence was seen between those aged 61-80 years. Figure 3 shows that the incidence increased with age but declined above the age of 80 years.

**Figure 3.**
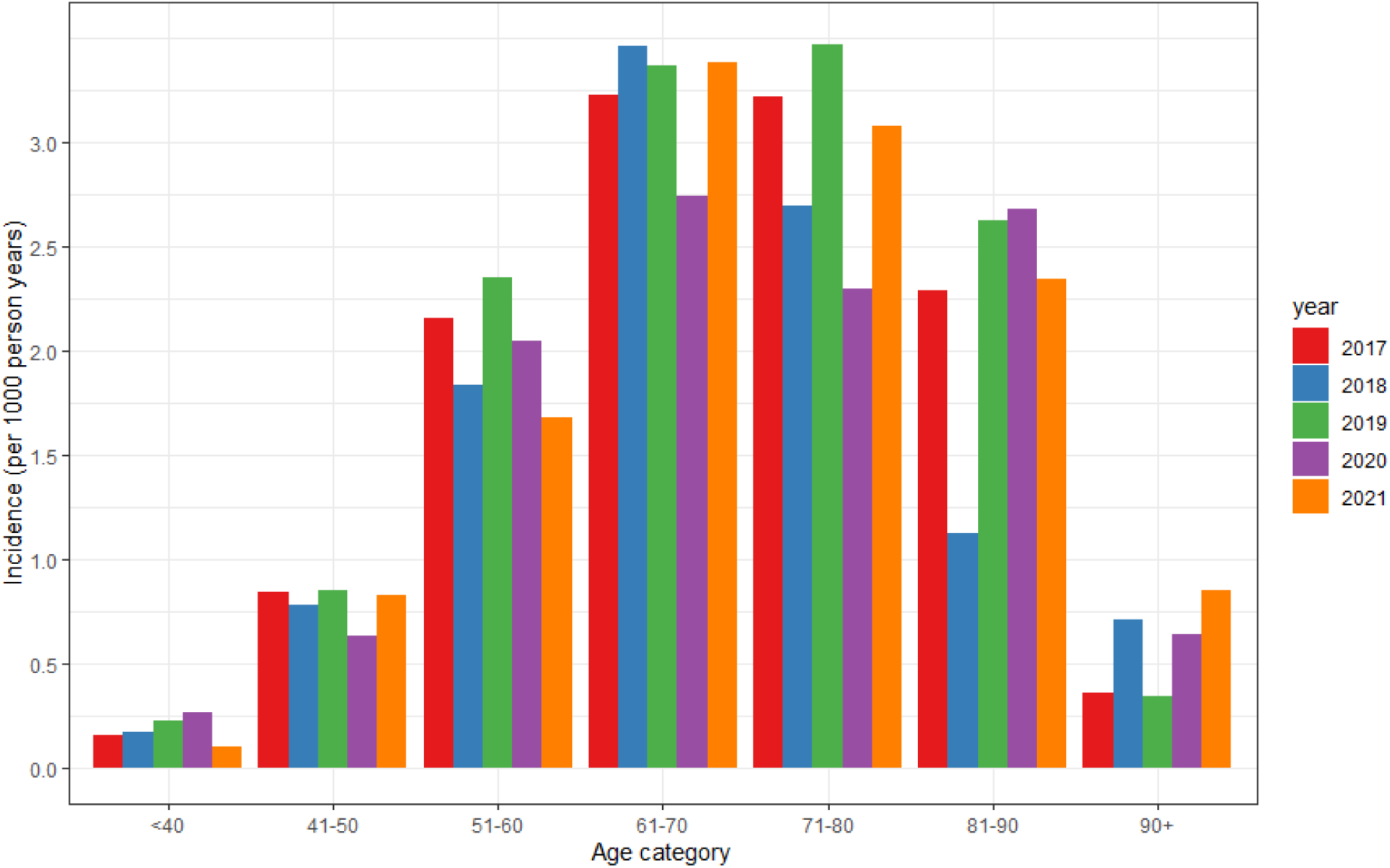
The incidence of Dupuytren’s disease in the AHON registry specified per year of observation and age category.

### Prevalence

The point prevalence of DD in 2021 was 1.99% (95% CI: 1.89-2.10). Figure 3 shows that the prevalence was the lowest under the age of 40, increasing with every age group to peak at 71-80 years for males and 81-90 years for females to decrease again after 90 years of age. The prevalence was higher for males in every age group: the overall prevalence for males was 2.50% (95%CI 2.33 – 2.67) compared to 1.50% (95%CI 1.38 – 1.64) for females.

**Figure 4.**
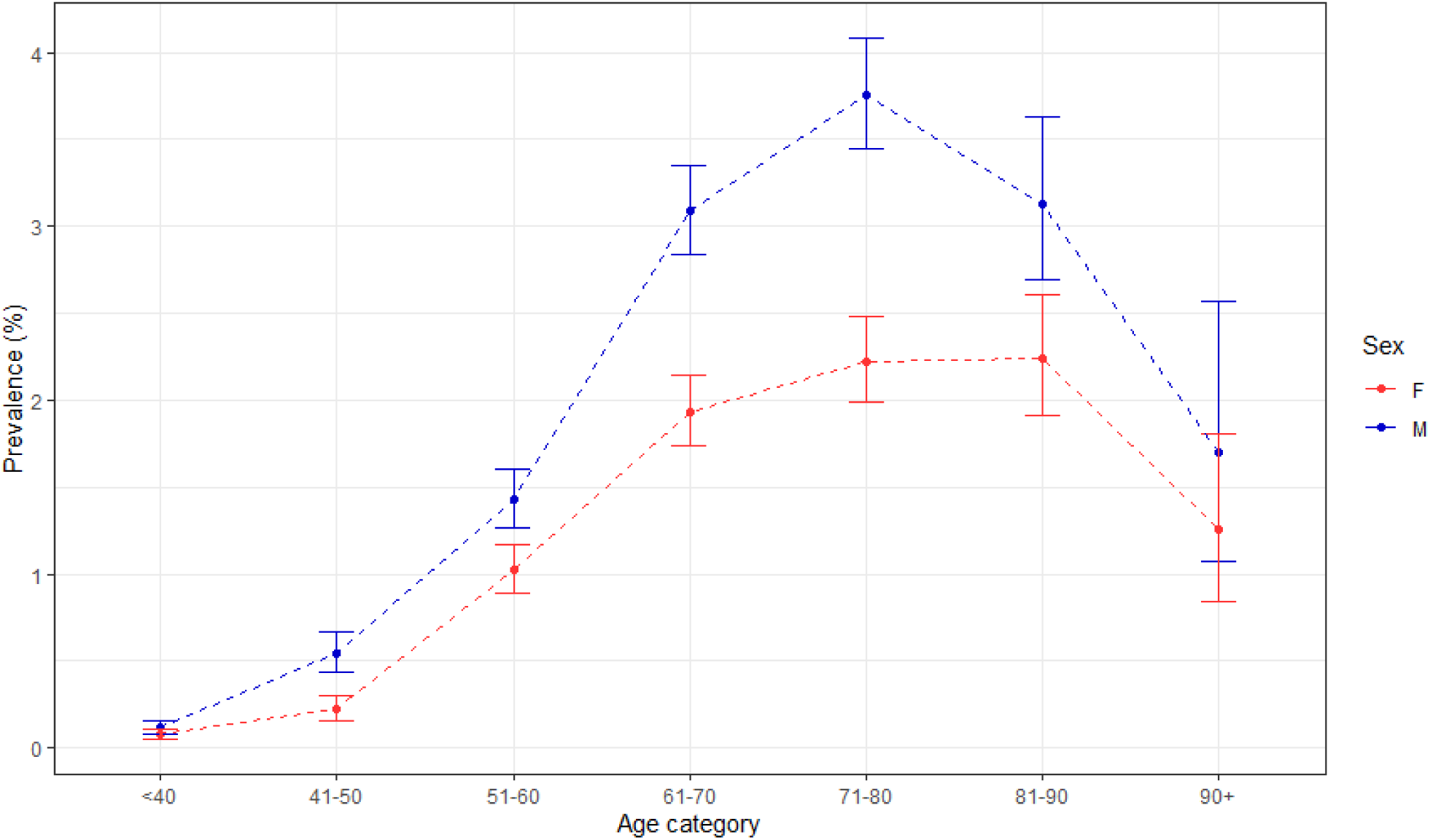
2021 mid-year prevalence of Dupuytren’s disease in the AHON registry per age category, stratified by sex.

## Discussion

Our study reports the incidence and prevalence of patients presenting with DD in primary care. The results show an incidence ranging between 1.41 to 1.72/1000 person years and an overall prevalence of 1.99%. Both incidence and prevalence are higher among males and increase with age, peaking between 61 to 80 years.

Our results show that DD is not a rare disease in primary care. In comparison, in 2019 the DD incidence was higher than the incidence of appendicitis or diverticulosis (both 1.6/1000 persons).(25)

Our reported incidence somewhat exceeds the incidence from a Swedish cohort study that described the number of people seeking medical care for DD in primary, secondary and tertiary care in 2013.(14) They reported annual incidences of “health-seeking patients with DD” of 2.7/1000 person years for males and 0.9/1000 person years for females ≥ 50 years. Their results of higher incidence rates in males peaking between ages 70-80 years, agrees with our results. They also reported a lower overall point-prevalence compared to our results of 0.92% in participants ≥20 years (1.35% in males and 0.50% in females). In line with our results, the highest prevalence was among those aged >70 for both males and females.(14)

A recent cohort study, originating from a primary care registration database in the United Kingdom, described an increasing DD incidence trend from 2000 to 2013, of 0.303/1000 person-years to 0.587/1000 person-years. They reported a point prevalence of 0.67% in 2013 which is lower compared to our findings.(26)) The fact that we found higher rates, might be explained by the different time window in which the studies were conducted and are in line with the finding of an increasing incidence of symptomatic DD.(27) Another explanation for the discrepancy might be the difference in ethnicity between the study populations. DD is particularly common in areas where people of Northern European descent live.(28) Our study was conducted in the Northern region of the Netherlands, which has a lower percentage of people with migration backgrounds compared to the United Kingdom(29,30). A final reason might be related to the different methodology of DD assessment. In the UK study, the diagnosis DD was only assessed via registrational coding, whereas we additionally employed a text-mining algorithm. The sole use of registrational coding for DD assessment might have led to an underestimation of the prevalence.(18)

Two studies reported a much higher prevalence.(12,31) These studies screened for signs of DD in the general population aged >50 years and reported point prevalences of 22.1% and 32%. In concordance with our results, males were more often affected and the prevalence peaked between the ages of 70-80 years. One of these studies was also conducted in the Northern region of the Netherlands.(12) Despite this, our study showed a much lower prevalence (22% vs. 2%). This difference can be explained by the fact that they looked for DD signs in a stratified random sample of the general population by visiting people at home, while our population consisted of people who sought medical care and then were diagnosed with DD by the GP. This illustrates that many people with DD are asymptomatic and that only a minority finds their symptoms serious enough to seek medical consultation. This may also account for the lower prevalence in the oldest age groups, as they might have a decreased tendency to seek medical help for DD.

The slight decrease in incidence in 2020 and 2021 in our study might be explained by the COVID-19 pandemic. During the pandemic, access to primary health care was restricted by national and local regulations. People were discouraged to visit their GP for not directly life-threatening symptoms or were afraid to catch a COVID-19 infection. (32)

A strength of our study is the use of a large primary care registration-based dataset, that consisted data of almost 300.000 GP registered persons. However, the primary goal of registration data is to monitor patient care and not to collect data for research purposes. This might have led to under registration of some of the descriptive information such as lifestyle factors.

Because a large part of the participating practices registered in 2017 and provided patient information from 5 years prior, we chose to calculate the point prevalence in 2021. We decided not to calculate the prevalence rates of prior years, because we wanted to maintain a run-in period of at least 10 years to minimize underestimation. However, we cannot exclude that the ‘first DD contact’ was preceded by a previous contact many years earlier, which might result in an overestimation of the incidence rate.

Another strength is that selection bias is unlikely, because the Netherlands uses a primary healthcare system in which all people are registered with a GP. However, the Northern region of the Netherlands is not representative of the Netherlands(30), which may make our results somewhat less generalisable for the whole country.

A final strength of our study is the use of a text-mining algorithm to extract DD diagnoses, compensating for the possible pitfalls in the use of ICPC codes (ICPC L99.03) among GPs and addressing the impracticality and potential errors associated with manually scoring large datasets. Our algorithm was validated through a three-step procedure and excellent agreement was achieved with manually scored data in a validation subset of the data.

## Conclusion

Based on GP codes and text mining of GP notes, we observed an incidence of 1.72/1000 person years and an overall prevalence of 1.99% of people with DD seeking medical help in a typical gatekeeping setting. Our results provide an insight into the relevant population of patients with symptomatic DD that seek medical care for the first time, which might – at least in part - be the future target group for potential disease controlling treatments.

## Declarations/ Acknowledgements

### Ethical approval

This study was approved by the local medical ethics committee (METc UMCG 202100077).

## Acknowledgements

This research was conducted using the AHON database. We are grateful for the support work provided by Feikje Groenhof and Ronald Wilmink.

## Data availability

The datasets used and/or analysed during the current study are available from the corresponding author and AHON commission on reasonable request.

## Contributorship

All authors contributed to the conceptualization and methodology of the study. The statistical analysis and data interpretation were performed by RvS, MB and DB. The manuscript was written by RvS and all authors provided feedback and commented on previous versions of the manuscript. All authors read and approved the final manuscript.

## References

1. Zyluk A, Jagielski W. The effect of the severity of the dupuytren’s contracture on the function of the hand before and after surgery. Journal of Hand Surgery. 2007;32(3):326–9.

2. Engstrand C, Krevers B, Nylander G, Kvist J. Hand function and quality of life before and after fasciectomy for dupuytren contracture. Journal of Hand Surgery [Internet]. 2014;39(7):1333–1343.e2. Available from: 10.1016/j.jhsa.2014.04.029

3. Reilly RM, Stern PJ, Goldfarb CA. A retrospective review of the management of Dupuytren’s nodules. Journal of Hand Surgery. 2005;30(5):1014–8.

4. Gudmundsson KG, Arngrímsson R, Jónsson T. Eighteen years follow-up study of the clinical manifestations and progression of Dupuytren’s disease. Scand J Rheumatol. 2001;30(1):31–4.

5. Van Den Berge BA, Werker PMN, Broekstra DC. Limited progression of subclinical Dupuytren’s disease results from a prospective cohort study. Bone and Joint Journal. 2021;103 B(4):704–10.

6. Van Rijssen AL, Ter Linden H, Werker PMN. Five-year results of a randomized clinical trial on treatment in Dupuytren’s disease: Percutaneous needle fasciotomy versus limited fasciectomy. Plast Reconstr Surg. 2012;129(2):469–77.

7. Ball C, Izadi D, Verjee LS, Chan J, Nanchahal J. Systematic review of non-surgical treatments for early dupuytren’s disease. BMC Musculoskeletal Dis. 2016;17(1):345.

8. Nanchahal J, Ball C, Rombach I, Williams L, Kenealy N, Dakin H, et al. Anti-tumour necrosis factor therapy for early-stage Dupuytren’s disease (RIDD): a phase 2b, randomised, double-blind, placebo-controlled trial. Lancet Rheumatol [Internet]. 2022;4:e407–16. Available from: 10.1016/S2665-9913(22)00093-5

9. Forrest CB. Primary care in the United States: Primary care gatekeeping and referrals: Effective filter or failed experiment? Br Med J. 2003;326(7391):692–5.

10. B. S, L. S. Policy relevant determinants of health: An international perspective. Health Policy (New York) [Internet]. 2002;60(3):201–18. Available from: http://ovidsp.ovid.com/ovidweb.cgi?T=JS&PAGE=reference&D=emed5&NEWS=N&AN=2002142554

11. van Dijk CE, Korevaar JC, Koopmans B, de Jong JD, de Bakker DH. The primary-secondary care interface: Does provision of more services in primary care reduce referrals to medical specialists? Health Policy (New York). 2014;118(1):48–55.

12. Lanting R, Van Den Heuvel ER, Westerink B, Werker PMN. Prevalence of Dupuytren disease in The Netherlands. Plast Reconstr Surg. 2013;132(2):394–403.

13. Hindocha S, McGrouther DA, Bayat A. Epidemiological evaluation of dupuytren’s disease incidence and prevalence rates in relation to etiology. Hand. 2009;4(3):256–69.

14. Nordenskjöld J, Englund M, Zhou C, Atroshi I. Prevalence and incidence of doctor-diagnosed Dupuytren’s disease: A population-based study. Journal of Hand Surgery: European Volume [Internet]. 2017;42(7):673–7. Available from: 10.1177/1753193416687914

15. Salari N, Heydari M, Hassanabadi M, Kazeminia M, Farshchian N, Niaparast M, et al. The worldwide prevalence of the Dupuytren disease: a comprehensive systematic review and meta-analysis. J Orthop Surg Res. 2020;15(1):1–13.

16. Lanting R, Broekstra DC, Werker PMN, van den Heuvel ER. A systematic review and meta-analysis on the prevalence of dupuytren disease in the general population of western countries. Plast Reconstr Surg. 2014;133(3):593–603.

17. Broekstra DC, Kuo RYL, Burn E, Pireto-Alhambra D, Furniss D. Dupuytren’s disease: Prevalence, incidence, and lifetime risk of surgical intervention A population-based cohort analysis. Plast Reconstr Surg. 2022;10.

18. Maret-Ouda J, Tao W, Wahlin K, Lagergren J. Nordic registry-based cohort studies: Possibilities and pitfalls when combining Nordic registry data. Scand J Public Health. 2017;45(17_suppl):14–9.

19. Twickler R, My B, Groenhof F, Sulim K, Ab E, Mh B, et al. Data Resource Profile: Registry of Electronic Health Records of General Practices in the North of the Netherlands (AHON). Int J Epidemiol. 2024;

20. WHO. Guidelines for ATC classification and DDD assignment. 2021.

21. Verbeke M, Schrans D, Deroose S, de Maeseneer J. The international classification of primary care (ICPC-2): An essential tool in the EPR of the GP. Stud Health Technol Inform. 2006;124(February):809–14.

22. Wickham H, Averick M, Bryan J, Chang W, McGowan L, François R, et al. Welcome to the tidyverse. J Open Source Softw [Internet]. 2019 Nov;4(43):1686. Available from: 10.21105/joss.01686

23. Pace M. Office for National Statistics. Revised European Standard Population [Internet]. 2013. Available from: https://ec.europa.eu/eurostat/documents/3859598/5926869/KS-RA-13-028-EN.PDF/e713fa79-1add-44e8-b23d-5e8fa09b3f8f

24. Aragon TJ. epitools: Epidemiology Tools [Internet]. 2020. Available from: https://cran.r-project.org/package=epitools

25. Heins M, Bes J, Weesie Y, Davids R, Winckers M, Korteweg L, et al. Zorg door de huisarts. Nivel zorgregistraties Eerste Lijn: jaarcijfers 2021 en trendcijfers 2017-2021. [Internet]. Nivel. 2022. 36 p. Available from: https://www.nivel.nl/sites/default/files/bestanden/1004116.pdf

26. Broekstra DC, Kuo RYL, Burn E, Prieto-Alhambra D, Furniss D. Dupuytren’s disease: Prevalence, incidence, and lifetime risk of surgical intervention A population-based cohort analysis. Plast Reconstr Surg. 2022 Nov;

27. Balachandran A, de Beer J, James KS, van Wissen L, Janssen F. Comparison of Population Aging in Europe and Asia Using a Time-Consistent and Comparative Aging Measure. J Aging Health. 2020;32(5–6):340–51.

28. Gudmundsson KG, Arngrímsson R, Sigfússon N, Björnsson Á, Jónsson T. Epidemiology of Dupuytren’s disease: Clinical, serological, and social assessment. The Reykjavik Study. J Clin Epidemiol. 2000;53(3):291–6.

29. Office for National Statistics. Regional ethnic diversity [Internet]. 2018. Available from: https://www.ethnicity-facts-figures.service.gov.uk/uk-population-by-ethnicity/national-and-regional-populations/regional-ethnic-diversity/latest

30. CBS. Hoeveel mensen met een migratieachtergrond wonen in Nederland? [Internet]. 2022. Available from: https://www.cbs.nl/nl-nl/dossier/dossier-asiel-migratie-en-integratie/hoeveel-mensen-met-een-migratieachtergrond-wonen-in-nederland-

31. Degreef I, de Smet L. A high prevalence of Dupuytren’s disease in Flanders. Acta Orthop Belg. 2010 Jun;76(3):316–20.

32. Lambooij MS, Heins M, Jansen L, Meijer M, Vader S, de Jong J. Avoiding GP care during the coronavirus pandemic. 2022. p. 1–73.

