## Supplementary data for "The incidence and prevalence of Dupuytren’s disease in primary care: Results from a text mining approach on registration data"

**Appendix 1:** **Removing/merging duplicate subjects.**

The following analysis was done in R (version 4.0.5) using packages tidyverse, irr, haven and foreign.

We received data including all patients since the start of the AHON in 1998, this included over 500.000 patient identification numbers (ID’s), which represented a patient. Each ID had two different subject identification pseudonyms, based on date of birth, gender and postal codes.

A patient identification number can have one of five different possible pseudonym combinations and based on the admission tarrifs (adm.tar.) the following actions may occur (*table S1).*

| *Table S1: Different pseudonym combinations* | | | | |
| --- | --- | --- | --- | --- |
|  | **Pseudonyms** | **Admission Tarrifs** | **Conclusion** | **Action** |
| 1 | Missing C pseudonym &  Missing N pseudonym | 1.1 No adm.tar.  1.2 Adm.tar. available | 1.1 No true ID  1.2 True ID | 1.1 Exclusion  1.2 No action |
| 2 | C pseudonym duplicate &  N Pseudonym duplicate | *Irrelevant* | Duplicate | Merge |
| 3 | Unique C pseudonym &  Duplicate N pseudonym  **Or vice versa** | 3.1 Only 1 pat-id has adm.tar.  3.2 Both pat-id mis adm.tar.  3.3 Two or more pat-id have adm.tar.:  3.3.1 Overlapping adm.tar. dates  3.3.2 Not overlapping adm.tar. dates | 3.1 Duplicate  3.2 Duplicate  3.3.1 Unique ID’s  3.3.2 Duplicate | 3.1 Merge  3.2 Merge  3.3.1 No action  3.3.2 Merge |
| 4 | Missing C pseudonym  Duplicate N pseudonym  **Or vice versa** | *Irrelevant* | Duplicate | Merge |
| 5 | Unique C pseudonym  Unique N pseudonym | *Irrelevant* | Unique ID | No action |

Before we started merging duplicates we added the variable ‘diagnosis DD’ to the dataframe (*appendix 2)*.

We started by creating a new ID variable (Dup-NC) which gave a duplicate ID number if

- the N pseudonym and the C pseudonym were duplicate (situation 2), or
- the N pseudonym was duplicate and the C pseudonym is missing (situation 4), or
- the N pseudonym is missing and the C pseudonym is duplicate (situation 4), or
- the N pseudonym is missing and the C pseudonym is missing (situation 1).

*my_df <- my_df %>% group_by(Pseudoniem_N, Pseudoniem_C) %>% mutate("Dup_NC" = cur_group_id())*

If the N pseudonym (Dup_N) or C pseudonym (Dup_C) was missing it had a value ‘NA’. We created two new variables which flagged ‘true’ if ‘Dup_N’ or ‘Dup_C’ was duplicate and there were overlapping adm.tar. periods:

*my_df %>%*

*group_by(Dup_N) %>%*

*mutate( overlapN=(lead(inschrijf_first)>inschrijf_first &*

*lead(inschrijf_first)<inschrijf_last)|*

*(lead(inschrijf_last)>inschrijf_first &*

*lead(inschrijf_last)<inschrijf_last) ) %>%*

*fill(overlap, .direction="downup") %>%*

*ungroup()*

*my_df %>%*

*group_by(Dup_C) %>%*

*mutate( overlapC=(lead(inschrijf_first)>inschrijf_first &*

*lead(inschrijf_first)<inschrijf_last)|*

*(lead(inschrijf_last)>inschrijf_first &*

*lead(inschrijf_last)<inschrijf_last) ) %>%*

*fill(overlap, .direction="downup") %>%*

*ungroup()*

After this we created two new ID variables, ‘newID_N’ and ‘newID_C’, which gave a duplicate number if pseudonym N or C is duplicate and the variable ‘overlapN’ or ‘overlapC’ is ‘false’:

- NewID_N = if pseudonym N is double/duplicate and overlapN(FALSE) the pat_id's get a duplicate number assigned
- NewID_C = if pseudonym C is double/duplicate and overlapC(FALSE) the pat_id's get a duplicate number assigned

*My_df$newID_N <- with(my_df, data.table::rleid(Dup_N + cumsum(overlapN)))*

*My_df$newID_C <- with(my_df, data.table::rleid(Dup_C + cumsum(overlapC)))*

Now we had three ID variables: Dup_NC, newID_N and newID_C. Based on these three variables we created a final ID variable in which all possible duplicates (see table above) were given duplicate numbers:

*My_df$ID_duplicate <- c(1,*

*cumsum( sapply(*

*seq_len(nrow(assign_DD1)-1),*

*function(x) {*

*!any(mapply(`-`, assign_DD1[x, -1, drop = T], assign_DD1[x + 1, -1, drop = T]) == 0, na.rm = T)*

*}*

*)*

*) + 1*

*)*

After this, we assigned the same ‘diagnosis DD’ value (true/false) if the ID_duplicate variable was duplicate:

*My_df %>%*

*group_by(ID_duplicate) %>%*

*mutate(xDD = any(as.logical(diagnosisDD)) %>% ifelse(is.na(.), FALSE, .))*

So now all possible situations for duplicates were assigned a duplicate ID_duplicate number and the diagnosis DD (true/false) was assigned to all duplicates.

First we excluded the ID’s which corresponded with situation 1.1:

Patient ID’s with a missing C pseudonym and N Pseudonym were assigned the value ‘1’ for ‘Dup_NC’.

Patient ID’s were excluded if ‘Dup_NC’ was ‘1’ and there were no admission tarrifs date:

*My_df[ !(My_df$Dup_NC == "1" & is.na(My_df$admissiontarrif1)), ]*

Then we merged the first-registration date and the last de-registration date for the ID_duplicates:

*My_df %>%*

*group_by(ID_duplicate) %>%*

*mutate(Registrationdate = min(Registrationdate), Deregistrationdate = max(Deregistrationdate))%>%*

*ungroup()*

After this we retained only the unique/distinct ID_duplicate rows:

*My_df <-*

*My_df %>%*

*distinct(ID_duplicate, .keep_all = TRUE)*

**Appendix 2: Diagnosis Dupuytren via text-mining in R.**

The following analysis were done in SPSS statistics 23 and R (version 4.0.5) using packages tidyverse, irr, haven and foreign. We received the following data from the AHON registry: patients identification numbers (ID’s), gender, age, registration date, deregistration date, admission tariffs dates and two subject identification pseudonyms. We also received the ICPC code, contact dates and the free text per contact for ICPC codes of L99.03 (Dupuytren) or contacts with the word ‘dup’ in the free text.

To determine whether a patient was diagnosed with DD we identified 3 possibilities:

1. When the ICPC code L99.03 was registered in the contact.
2. When Dupuytren (or a incorrectly written version) was mentioned in the free-text.
3. When the ICPC code L99.03 was registered in the contact and DD was mentioned in the free-text.

To determine whether a patient had been diagnosed with DD, we created five variables: free-text DD, ICPC DD, suspected DD, no DD and suspected Ledderhose. These variables were created by several other variables that coded for different letter and word combinations in the free-text column and the ICPC column.

1) Free-text DD:

*dataframe$Dupu<-grepl("[Dd]upu", dataframe $tekst)*

*dataframe $Dupy<-grepl("[Dd]upy", dataframe $tekst)*

*dataframe $Dupi<-grepl("[Dd]upi", dataframe $tekst)*

*dataframe $Dupr<-grepl("[Dd]upr", dataframe $tekst)*

*dataframe <- mutate(dataframe, freetekst_DD= ifelse(*

*Dupu == "TRUE"|*

*Dupi == "TRUE"|*

*Dupy == "TRUE"|*

*Dupr == "TRUE", "TRUE", "FALSE"*

*))*

2) ICPC DD:

*dataframe$ICPC_DD<-grepl("L99.03", dataframe$icpc)*

3) Suspected DD:

For this variable we created a total of 119 letter and word combinations to filter out possible or suspected DD diagnosis. Below is a short example of the possible combinations.

*dataframe$verdDupu<-grepl("suspected [Dd]upu", dataframe$tekst)*

*dataframe$verdDupu1<-grepl("suspected [Dd]upi", dataframe$tekst)*

*dataframe$verdDupu2<-grepl("suspected [Dd]upy", dataframe$tekst)*

*dataframe$verdDupu3<-grepl("suspected [Dd]upr", dataframe$tekst)*

*dataframe$verdDupu4<-grepl("possible [Dd]upu", dataframe$tekst)*

*dataframe$verdDupu5<-grepl("possible [Dd]upi", dataframe$tekst)*

*dataframe$verdDupu6<-grepl("possible [Dd]upy", dataframe$tekst)*

*dataframe$verdDupu7<-grepl("possible [Dd]upr", dataframe$tekst)*

*dataframe <- mutate(dataframe, suspected_DD= ifelse(*

*verdDupu == "TRUE"|*

*verdDupu1 == "TRUE"|*

*verdDupu2 == "TRUE"|*

*verdDupu3 == "TRUE"|*

*verdDupu4 == "TRUE"|*

*verdDupu5 == "TRUE"|*

*verdDupu6 == "TRUE"|*

*verdDupu7 == "TRUE"* *, "TRUE", "FALSE"*

*))*

4) No DD:

For this variable we also first created 128 different letter and word combinations to filter out the contacts that did not involve a DD diagnosis. Below is a small example of possible combinations.

*dataframe $noDup<-grepl("no [Dd]upu", dataframe $tekst)*

*dataframe $noDup1<-grepl("no [Dd]upi", dataframe $tekst)*

*dataframe $noDup2<-grepl("no [Dd]upy", dataframe $tekst)*

*dataframe $noDup3<-grepl("no [Dd]upr", dataframe $tekst)*

*dataframe <- mutate(dataframe, no_DD= ifelse(*

*noDup == "TRUE"|*

*noDup1== "TRUE"|*

*noDup2 == "TRUE"|*

*noDup3 == "TRUE", "TRUE", "FALSE"*

*))*

5) Suspected Ledderhose

Because Ledderhose disease falls under the same ICPC code (L99.03) as DD, we created this variable to filter out the possible Ledderhose diagnosis (without simultaneous DD diagnosis). For this we created 8 different letter and word combinations, below an example of possible combinations.

*dataframe <- mutate(dataframe, suspected_Ledderhose= ifelse(*

*free-tekst_DD == "TRUE" &*

*ICPC_DD == "FALSE" &*

*(grepl("[Ll]edderhose", dataframe $tekst)|*

*grepl("[Ll]ederhose", dataframe $tekst)|*

*grepl("[Ll]ederhoz", dataframe $tekst)|*

*grepl("[Ll]edderhoz", dataframe $tekst), "TRUE", "FALSE"*

*))*

So each new variable included many different written versions and typo’s in the grepl function.

Finally we created the variable diagnosis DD with the mutate, grepl and ifelse function:

*Dataframe <- mutate(dataframe, diagnosis_DD = ifelse(*

*free-tekst_DD == “TRUE” |*

*ICPC_DD == “TRUE” &*

*suspected_DD == “FALSE” &*

*no_DD == “FALSE” &*

*suspected_Ledderhose == “FALSE”, “TRUE”, “FALSE”))*

**Appendix 3:** The RECORD statement – checklist of items, extended from the STROBE statement, that should be reported in observational studies using routinely collected health data.

|  | **Item No.** | **STROBE items** | **Location in manuscript where items are reported** | **RECORD items** | **Location in manuscript**  **where items are reported** |
| --- | --- | --- | --- | --- | --- |
| **Title and abstract** | | | | | |
|  | 1 | (a) Indicate the study’s design with a commonly used term in the title or the abstract (b) Provide in the abstract an informative and balanced summary of what was done and what was found | (a) page 1  (b) page 2 | RECORD 1.1: The type of data used should be specified in the title or abstract. When possible, the name of the databases used should be included.  RECORD 1.2: If applicable, the geographic region and timeframe within which the study took place should be reported in the title or abstract.  RECORD 1.3: If linkage between databases was conducted for the study, this should be clearly stated in the title  or abstract. | Page 2 |
| **Introduction** | | | | | |
| Background rationale | 2 | Explain the scientific background and rationale for the  investigation being reported | Page 3 |  |  |
| Objectives | 3 | State specific objectives, including any prespecified  hypotheses | Page 3,4 |  |  |
| **Methods** | | | | | |
| Study Design | 4 | Present key elements of study design early in the paper | Page 5 |  |  |
| Setting | 5 | Describe the setting, locations, and relevant dates, including  periods of recruitment, exposure, follow-up, and data collection | Page 5 |  |  |
| Participants | 6 | (a) *Cohort study* - Give the eligibility criteria, and the sources and methods of selection of participants. Describe methods of follow-up   *Case-control study* - Give the eligibility criteria, and the sources and methods of case ascertainment and control selection. Give the rationale for the choice of cases and controls *Cross-sectional study* - Give the eligibility criteria, and the sources and methods of selection of participants   (b) *Cohort study* - For matched studies, give matching criteria and number of exposed and unexposed   *Case-control study* - For matched studies, give matching  criteria and the number of controls per case | Page 5,6 | RECORD 6.1: The methods of study population selection (such as codes or algorithms used to identify subjects) should be listed in detail. If this is not possible, an explanation should be provided.  RECORD 6.2: Any validation studies of the codes or algorithms used to select the population should be referenced. If validation was conducted for this study and not published elsewhere, detailed methods and results should be provided.  RECORD 6.3: If the study involved linkage of databases, consider use of a flow diagram or other graphical display to demonstrate the data linkage process, including the number of individuals with linked data at each stage. | Page 5,6  Page 20,21 |
| Variables | 7 | Clearly define all outcomes, exposures, predictors, potential confounders, and effect modifiers. Give diagnostic criteria, if applicable. | Page 6,7 | RECORD 7.1: A complete list of codes and algorithms used to classify exposures, outcomes, confounders, and effect modifiers should be provided. If these cannot be reported, an  explanation should be provided. | Page 6,7  Page 22,23 |
| Data sources/ measurement | 8 | For each variable of interest, give sources of data and details of methods of assessment (measurement).  Describe comparability of  assessment methods if there is more than one group | Page 5,6 |  |  |
| Bias | 9 | Describe any efforts to address potential sources of bias | Page 5,6 |  |  |
| Study size | 10 | Explain how the study size was  arrived at | Page 5,6 |  |  |
| Quantitative variables | 11 | Explain how quantitative variables were handled in the analyses. If applicable, describe which groupings were chosen,  and why | Page 6,7 |  |  |
| Statistical methods | 12 | (a) Describe all statistical methods, including those used to control for confounding (b) Describe any methods used to examine subgroups and interactions (c) Explain how missing data were addressed (d) *Cohort study* - If applicable, explain how loss to follow-up was addressed   *Case-control study* - If applicable, explain how matching of cases and controls was addressed  *Cross-sectional study* - If applicable, describe analytical methods taking account of sampling strategy   (e) Describe any sensitivity   analyses | Page 7 |  |  |
| Data access and cleaning methods |  | .. |  | RECORD 12.1: Authors should describe the extent to which the investigators had access to the database population used to create the study population. | Page 5, 13 |
|  |  |  |  | RECORD 12.2: Authors should provide information on the data cleaning methods used in the study. | Page 20, 21 |
| Linkage |  | .. |  | RECORD 12.3: State whether the study included person-level, institutional-level, or other data linkage across two or more databases. The methods of linkage and methods of  linkage quality evaluation should be provided. | - |
| **Results** |  |  |  |  |  |
| Participants | 13 | (a) Report the numbers of individuals at each stage of the study (*e.g.*, numbers potentially eligible, examined for eligibility, confirmed eligible, included in the study, completing follow-up, and analysed) (b) Give reasons for non- participation at each stage.   Consider use of a flow diagram | Page 8 | RECORD 13.1: Describe in detail the selection of the persons included in the study (*i.e.,* study population selection) including filtering based on data quality, data availability and linkage. The selection of included persons can be described in the text and/or by means of the study flow diagram. |  |
| Descriptive data | 14 | (a) Give characteristics of study participants (*e.g.*, demographic, clinical, social) and information on exposures and potential confounders (b) Indicate the number of participants with missing data for each variable of interest (c) *Cohort study* - summarise follow-up time (*e.g.*, average and   total amount) | Page 8 |  |  |
| Outcome data | 15 | *Cohort study* - Report numbers of outcome events or summary measures over time  *Case-control study* – Report numbers in each exposure category, or summary measures of exposure  *Cross-sectional study* - Report  numbers of outcome events or summary measures | Page 8 |  |  |
| Main results | 16 | (a) Give unadjusted estimates and, if applicable, confounder- adjusted estimates and their precision (e.g., 95% confidence interval). Make clear which confounders were adjusted for and why they were included (b) Report category boundaries when continuous variables were categorized (c) If relevant, consider translating estimates of relative risk into absolute risk for a   meaningful time period | Page 8 |  |  |
| Other analyses | 17 | Report other analyses done— e.g., analyses of subgroups and interactions, and sensitivity  analyses | Page 8 |  |  |
| **Discussion** |  |  |  |  |  |
| Key results | 18 | Summarise key results with reference to study objectives | Page 10 |  |  |
| Limitations | 19 | Discuss limitations of the study, taking into account sources of potential bias or imprecision.  Discuss both direction and magnitude of any potential bias | Page 11 | RECORD 19.1: Discuss the implications of using data that were not created or collected to answer the specific research question(s). Include discussion of misclassification bias, unmeasured confounding, missing data, and changing eligibility over time, as they pertain to the study being  reported. | Page 11 |
| Interpretation | 20 | Give a cautious overall interpretation of results considering objectives, limitations, multiplicity of analyses, results from similar studies, and other relevant  evidence | Page 10,11,12 |  |  |
| Generalisability | 21 | Discuss the generalisability (external validity) of the study results | Page 11 |  |  |
| **Other Information** |  |  |  |  |  |
| Funding | 22 | Give the source of funding and the role of the funders for the present study and, if applicable, for the original study on which  the present article is based | Page 13 |  |  |
| Accessibility of protocol, raw data, and programming  code |  | .. |  | RECORD 22.1: Authors should provide information on how to access any supplemental information such as the study protocol, raw data, or  programming code. | Page 13 |

*Reference: Benchimol EI, Smeeth L, Guttmann A, Harron K, Moher D, Petersen I, Sørensen HT, von Elm E, Langan SM, the RECORD Working Committee. The REporting of studies Conducted using Observational Routinely-collected health Data (RECORD) Statement. *PLoS Medicine* 2015; in press.

*Checklist is protected under Creative Commons Attribution ([CC BY](http://creativecommons.org/licenses/by/4.0/)) license.
